# Optimising Remote Consulting and Home Assessment of Medically Vulnerable Rural Patients During Unscheduled and Planned Primary Care: Assessing the Feasibility of ORCHARD Intervention -A Feasibility Study

**DOI:** 10.64898/2026.04.08.26350378

**Authors:** Peter Murchie, Rosalind Adam, Shahana A Naqvi, Maria Ntessalean

## Abstract

**Background:** The COVID-19 pandemic significantly accelerated the adoption of telemedicine, but it also exposed gaps in effective remote clinical assessment, particularly for medically vulnerable patients in rural areas. The ORCHARD intervention aimed to address this by providing patients with a Medical Self-Assessment Box to enable self-reporting of vital signs during remote consultations.

**Methods:** A single-centre randomised mixed-methods feasibility trial recruited medically vulnerable patients from a rural general practice in Northeast Scotland. Participants in intervention group received a home medical equipment box for use during telemedicine consultations over six months. Patients and GPs were interviewed and transcripts were analysed using Framework Analysis.

**Results:** Twelve (15%) of 82 eligible invited patients enrolled. Six each were allocated to intervention and control group. 50%(n=3)patients in intervention group used equipment in 45%(5 of 11)teleconsultations and rated it helpful in all 5 uses (100%). The intervention group had 18% fewer primary care contacts than controls. All remote consultations were by telephone. Framework Analysis of patient interviews identified facilitators such as ease of use, improved triage access, reassurance, and barriers related to GP non-engagement and written instructions. GP interviews identified clinical value in patient-generated readings, alongside concerns regarding workload and patient over-monitoring.

**Conclusions:** Half of intervention participants used the medical-equipment box during remote consultations, all finding it useful, though frequency of use varied among particpants.A randomised controlled trial to evaluate the effectiveness of the Medical Self-Assessment Box for optimising remote consulting in medically vulnerable rural patients is feasible.Prior to a definitive trial refinements are recommended to patient labelling, GP engagement, and training materials.

## 1. Introduction

The COVID-19 pandemic accelerated the rapid adoption of remote consulting across primary care, including for patients considered medically vulnerable [1]. In Scotland, this group was formally identified by general practices early in the pandemic, with many advised to minimise face-to-face contact with health services for prolonged periods. While remote consulting enabled primary care services to maintain access during this period, it also highlighted longstanding challenges faced by rural populations in accessing timely healthcare.

Rural residents in Scotland experience structural barriers to primary and specialist care, including geographic isolation, travel burdens, and reduced availability of specialist services. These factors contribute to measurable health inequalities. Evidence suggests that rural cancer patients in Northeast Scotland experience poorer outcomes than their urban counterparts [2]. Findings from the Scottish National Cancer Diagnosis Audit indicate that rural patients may require more GP consultations and diagnostic investigations prior to referral and may follow different diagnostic pathways compared with urban patients. For certain cancers, including lung cancer, longer diagnostic intervals have been observed in rural areas [8,9]. More broadly, research in Scotland and internationally highlights the heterogeneity of rural populations, who experience varying levels of long-term illness and differing degrees of access constraints [10–12]. Within this context, telemedicine has been proposed as one mechanism for improving access to care, reducing travel requirements, and supporting continuity of services for rural patients [3,4].

However, remote consulting introduces important limitations for clinical assessment. Telephone or video consultations often lack access to basic physiological measurements that normally inform decision-making in face-to-face encounters, such as temperature, blood pressure, oxygen saturation, and peak flow. The absence of such data can create uncertainty in clinical assessment and may limit clinicians’ ability to safely triage, monitor symptoms, or identify early deterioration. This limitation may be particularly relevant for medically vulnerable rural patients, where geographic isolation can delay access to urgent in-person care.

Providing patients with simple home monitoring equipment could potentially strengthen remote consultations by enabling the collection of objective physiological data during or prior to teleconsultations. Access to contemporaneous measurements may support more structured clinical assessment, facilitate monitoring of symptom progression, and provide additional reassurance for both patients and clinicians when deciding whether further investigation or face-to-face review is required. In this way, home monitoring may improve processes of care within telemedicine by reducing diagnostic uncertainty and enabling more informed clinical decision-making.

Emerging evidence supports this potential role. Studies conducted during and after the pandemic indicate that telemedicine can improve access equity and reduce avoidable travel for rural patients [3,4]. Evidence from UK primary care also suggests that home self-monitoring tools can enhance remote management of conditions such as hypertension and respiratory disease and can support safer decision-making in teleconsultations [14,15]. However, the integration of monitoring technologies into routine care involves more than simply providing equipment. Greenhalgh and colleagues have emphasised that effective remote care depends on a wider sociotechnical system in which patients, clinicians, technologies, and organisational routines interact [13]. Even relatively simple monitoring devices must be incorporated into complex healthcare systems, requiring patients to collect and communicate measurements, clinicians to interpret them appropriately, and practices to adapt workflows accordingly.

Recent national policy developments in Scotland further emphasise the role of remote monitoring in future healthcare delivery. Initiatives such as Connect Me remote-monitoring services, the Digital Front Door programme, and Digital Hospital @ Home models highlight the increasing use of home-based monitoring technologies to support accessible and sustainable care [16]. Despite this policy momentum, important questions remain about how such tools can be integrated into routine primary care. In particular, there is limited evidence regarding the feasibility, acceptability, and practical use of home monitoring for medically vulnerable patients living in rural areas.

The ORCHARD feasibility study was developed in response to this evidence gap. The study explored whether providing medically vulnerable rural patients with a Medical Self-Assessment Box containing simple physiological monitoring equipment could enhance remote consultations with their general practitioners. The intervention aimed to support teleconsultations by enabling patients to collect and share basic physiological measurements that would otherwise be unavailable during remote assessment.

Given the complexity of integrating even simple monitoring tools into routine care, the study adopted a mixed-methods approach to explore both practical feasibility and user experience. This paper reports findings from this pilot feasibility study, combining preliminary quantitative data with qualitative interviews conducted with participating patients and clinicians. The study aimed to:

1. Explore whether medically vulnerable rural patients are willing to receive and use a Medical Self-Assessment Box.
2. Examine how patients used the equipment in the context of telemedicine consultations.
3. Identify implications for refining the intervention and informing the design of a future UK-wide evaluation.

Together, these findings contribute to the emerging evidence base on remote monitoring in primary care and provide early insights into how telemedicine models may be adapted to better support medically vulnerable rural populations.

## 2. Methods

### Study Design

The ORCHARD study was a single-centre, randomised parallel-group feasibility trial with an embedded qualitative component, conducted over six months at a rural general practice in Northeast Scotland. Participants were randomised 1:1 to receive either a Medical Self-Assessment Box in addition to usual care (intervention group) or usual care alone (control group). The primary purpose of the study was to assess the feasibility and acceptability of implementing the intervention within routine rural primary care rather than to evaluate clinical effectiveness.

Feasibility was considered in relation to several practical aspects of trial delivery and intervention use, including the ability to recruit and retain medically vulnerable rural patients, the willingness of participants to receive and use the Medical Self-Assessment Box, and the extent to which physiological measurements could be incorporated into remote consultations with general practitioners. These indicators were used to inform whether the intervention and study procedures could be implemented in their current form, required modification, or would present significant barriers to wider evaluation.

Quantitative feasibility data were collected to examine aspects such as participant recruitment and retention, intervention uptake, patterns of equipment use, and potential changes in healthcare utilisation during the study period. In parallel, semi-structured qualitative interviews were conducted with participating patients and clinicians. These interviews were designed to provide detailed insights into the intervention mechanisms and implementation processes, including acceptability, usability of the equipment, perceived impact on remote consultations, and barriers or facilitators to integration within routine primary care

### Setting and Participants

The study was conducted at a single rural general practice in Northeast Scotland, serving a geographically dispersed population across Scottish Government Urban–Rural Classification categories 3–6. Ethical approval was granted by the relevant Research Ethics Committee (REC reference: 22/SW/0151).

Potential participants were identified by searching the electronic health record (eHR) for two criteria: residence in a rural postcode (Urban–Rural Classification categories 3–6) and designation by the practice as shielding during the COVID-19 pandemic. Patients with complex mental health or complex social health needs were excluded after screening by a General Practitioner within their practice. Ninety patients were identified as potentially eligible, of whom eight were excluded on these grounds, leaving 82 patients who were sent postal invitation packs. Those who expressed interest were contacted by the research fellow (MN) to discuss the study and obtain written informed consent. Twelve patients (15%) consented to participate and were enrolled. The study aimed to recruit 40 participants (20 per group); this target was not achieved due to recruitment challenges in the post-pandemic clinical environment , lack of sending second invitation and some confusion regarding shielding status.• MN remotely accessed the eHR and collected baseline clinical and demographic data from records of consenting patients.•MN remotely accessed the eHR once per week for 6 months to determine if any of the patients had had a consultation. If they had consulted, MN recorded relevant details in RedCap software.

All patient participants were invited to participate in an end of study telephone interview with a researcher (SN). All GPs at the participating practice were also invited by email to participate in an end of study telephone interview . those who consented were contacted by telephone for an interview arrangement. GPs were invited by email ,those who consented were contacted by telephone for an interview. GP surgery received reimbursement of £264 for reviewing the list of 200 participants. The practice were also reimbursed £80 for each GP interview(x 4) conducted with the research team

### Randomisation and Allocation

Following completion of baseline questionnaires, consenting participants were randomised 1:1 to the intervention group or the control group using an online randomisation tool (www.randomizer.com). Allocation was communicated to participants by post. Given the nature of the intervention, blinding of participants or clinicians to allocation was not possible. As this was a feasibility study with a small target sample, the randomisation process itself was assessed as a feasibility outcome, and perfect balance between groups was not expected or required.

### Intervention

Participants allocated to the intervention group received a Medical Self-Assessment Box containing four devices: a pulse oximeter, a digital sphygmomanometer, a tympanic thermometer, and a peak-flow meter. Each box was accompanied by brief illustrated written instructions to support independent use. Equipment was primarily intended for use during remote consultations to enable participants to self-report vital sign readings to their clinician in real time, though participants were not restricted from self-monitoring between consultations.

Electronic health records were flagged to alert both in-hours and out-of-hours clinicians to the participant’s equipment holder status, though in practice this flagging was not consistently implemented, which is acknowledged as a study limitation.

Participants remained under the care of their usual GP throughout and received no additional clinical input beyond that described above.

### Control Group

Participants allocated to the control group received usual primary care for the duration of the six-month study period. No equipment, additional consultations, or study-specific materials were provided to this group.

### Study Measures

Feasibility outcomes were the primary focus of data collection, organised across four domains:

#### Recruitment and Retention

Feasibility of recruitment was assessed by the proportion of invited patients who consented to participate and the proportion who completed the six-month follow-up period. Withdrawal rates and reasons for non-participation were recorded where available.

#### Intervention Uptake and Equipment Use

Equipment use was recorded both per participant and per remote consultation over the six-month study period. Use of equipment and perceived usefulness was captured via a brief post-consultation questionnaire completed by the patient following any telemedicine contact in which the equipment was used.

#### Clinical and Service Use Outcomes

Electronic medical records were reviewed at baseline and six months to extract: total number of primary care contacts, mode of consultation (face-to-face, telephone, or video), unscheduled out-of-hours contacts, hospital admissions, and survival status at six months. These data were collected for both groups to allow descriptive comparison.

#### Patient-Reported Outcome Measures

All participants completed the SF-12 (measuring physical and mental health-related quality of life) and the PHQ-4 (a four-item measure of anxiety and depression) at baseline.Due to change in researcher(MN) role above data was not be collected at end of 6 months.. Baseline demographic data were also collected, including age, gender, and prior ownership of home medical equipment (digital thermometer, blood pressure monitor, pulse oximeter, and spirometer).

#### Qualitative Outcomes

Semi-structured interviews were conducted with a purposive sample of patients and with GPs from the study practice. Patient interviews explored experiences of equipment use, ease of use, perceived value, and barriers to use. GP interviews explored clinical utility of patient-generated readings, impact on remote decision-making, workload implications, and views on wider implementation. Interviews were audio-recorded, transcribed verbatim, and analysed using the five-stage Framework Analysis approach: familiarisation with the data, identifying a thematic framework, indexing, charting, and mapping and interpretation.

### Data Management and Analysis

#### Quantitative Analysis

All participant data were anonymised at the point of collection and stored securely in accordance with University of Aberdeen data management protocols and NHS Grampian information governance requirements. Electronic health record data were extracted at baseline and during six months from Vision using RedCap by MN and recorded in patient files in RedCap. Quantitative feasibility and clinical data were summarised using descriptive statistics. Continuous variables are reported as means with standard deviations or medians with interquartile ranges as appropriate. Categorical variables are reported as frequencies and percentages. No inferential statistical testing was pre-specified, in keeping with the descriptive aims of the study. All quantitative data were analysed using. Quantitative data was analyzed by SN.

#### Qualitative Analysis

Semi-structured interviews with patients and GPs were conducted by SN. Qualitative interview data were organised using NVivo and analysed using Framework Analysis._Analysis was led by SN, with an independent review of a sample of transcripts by RA to ensure analytical rigour. Disagreements were resolved through discussion and consensus. As practising GPs, SN and RA , additionally RA with qualitative research experience brought relevant clinical perspective to the analysis, which was acknowledged as part of the reflexive approach to qualitative data interpretation.

#### Mixed Methods Integration

Quantitative and qualitative findings were integrated using a convergent mixed-methods approach. Qualitative data were used to explain and contextualise patterns observed in the quantitative results, particularly regarding variability in equipment use, non-use in specific clinical contexts such as palliative care, and differences in consultation rates between groups.

## 4. Results

### Recruitment and Retention

Ninety patients were screened for eligibility by AC, of whom eight were excluded by their general practice due to complex mental or social health needs. Out of the 82 eligible patients who were sent postal invitations, 12 (15%) returned a signed consent form and were subsequently enrolled. All 12 participants completed the six-month follow-up period, indicating a 100% retention rate. No participants withdrew during the study. Three invited patients who were not aware of their shielding code declined the invitation after receiving clarification.

Six participants were allocated to the intervention group and six to the control group. All recruited participants accepted randomisation, and no participants declined allocation to the intervention group. Baseline questionnaire completion was 100% (n=12). Pre study PHQ-4 and SF-12 questionnaire completion rates were 100% However , post study .PHQ SF12 completion were not requested.

### Baseline Characteristics

The baseline characteristics of the participants are summarized in Table 1. The mean age was 65.4 years (SD 11.3), with a range of 50 to 84 years. The intervention group was younger on average (mean 61.3, SD 9.8) than the control group (mean 69.5, SD 12.1). Five participants (42%) were female and seven (58%) were male. All participants had complex multimorbidity consistent with the medically vulnerable designation, including cancer, chronic respiratory, cardiovascular, and autoimmune conditions.

**Table 1.** Baseline characteristics of participants.

| Characteristic | Intervention Group (n=6) | Control Group (n=6) |
| --- | --- | --- |
| Age, mean (SD) | 61.3 (9.8) | 69.5 (12.1) |
| Age range | 52–80 years | 50–84 years |
| Gender: Female, n (%) | 3 (50%) | 2 (33%) |
| Gender: Male, n (%) | 3 (50%) | 4 (67%) |
| Questionnaire preference: Online | 5 (83%) | 6 (100%) |
| PHQ-4 total score, mean (SD) | 3.2 (3.6) | 2.7 (3.0) |
| PHQ-4 anxiety subscale, mean (SD) | 2.0 (2.3) | 1.7 (2.0) |
| PHQ-4 depression subscale, mean (SD) | 1.2 (2.4) | 1.0 (1.1) |
| PHQ-4 score $\geq 3$ (clinically significant), n (%) | 2 (33%) | 3 (50%) |
| SF-12 PCS raw score, mean (SD) | 15.5 (4.5) | 14.3 (3.4) |
| SF-12 MCS raw score, mean (SD) | 19.3 (4.5) | 21.2 (3.8) |
| General health: Excellent/Very good, n (%) | 2 (33%) | 1 (17%) |
| General health: Good, n (%) | 2 (33%) | 3 (50%) |
| General health: Fair/Poor, n (%) | 2 (33%) | 2 (33%) |
| BP monitor already owned, n (%) 7 | 2(28%) | 5(71%) |
| Thermometer already owned, n (%)4 | 2(50%) | 2(50%) |
| Pulse oximeter already owned, n (%)2 | 2(100%) | 0 |
| Spirometer already owned, n(%) | 0 | 0 |

At baseline, 7/12 participants (58%) already owned a blood pressure monitor, 4/12 (33%) owned a digital thermometer, 2/12 (17%) owned a pulse oximeter, and no participants owned a spirometer. The majority preferred to complete questionnaires online (11/12, 92%).

PHQ-4 scores at baseline were broadly comparable between groups (intervention mean 3.2, SD 3.6; control mean 2.7, SD 3.0). Two intervention participants (33%) and three control participants (50%) scored ≥3, indicating clinically significant anxiety or depression. SF-12 physical and mental component scores were broadly similar between groups, though the intervention group reported slightly lower general health on average.

### Service Use

Table 2 shows the service use data over the six-month study period. The intervention group had 25 total primary care contacts compared to 36 in the control group, representing 31% fewer contacts overall. In the intervention group, 14 contacts (56%) were face-to-face and 11 (44%) were by telephone. All remote consultations across both groups were conducted by telephone; no video consultations occurred during the study period.

**Table 2.** Service use over six months.

| Outcome | Intervention Group (n=6) | Control Group (n=6) |
| --- | --- | --- |
| Total primary care contacts, n | 25 | 36 |
| Face-to-face / house visit, n (%) | 14 (56%) | 17 (47%) |
| Telephone consultations, n (%) | 11 (44%) | 20 (56%) |
| Video consultations, n (%) | 0 (0%) | 0 (0%) |
| Hospital admissions (total), n | 0 | 0 |
| Emergency hospital admissions, n | 0 | 0 |
| Ambulance call-outs, n | 0 | 0 |
| Survival at 6 months, n | 5 | 6 |

There were no hospital admissions, emergency admissions, or ambulance call-outs in either group during the study period. One participant in the intervention group died before the six-month follow-up, leaving five survivors in this group compared to six in the control group. The intervention group participant with the highest number of contacts (n=11, 44% of all intervention contacts) was a palliative care patient.

### Intervention Uptake and Equipment Use

The equipment usage patterns of the six intervention group participants are shown in Table 3. Out of six participants, three (50%) used the Medical Self-Assessment Box during at least one remote consultation. Equipment was used in 5 of 11 (45%) telephone consultations in the intervention group and was rated helpful in all five uses (100%).

**Table 3.**
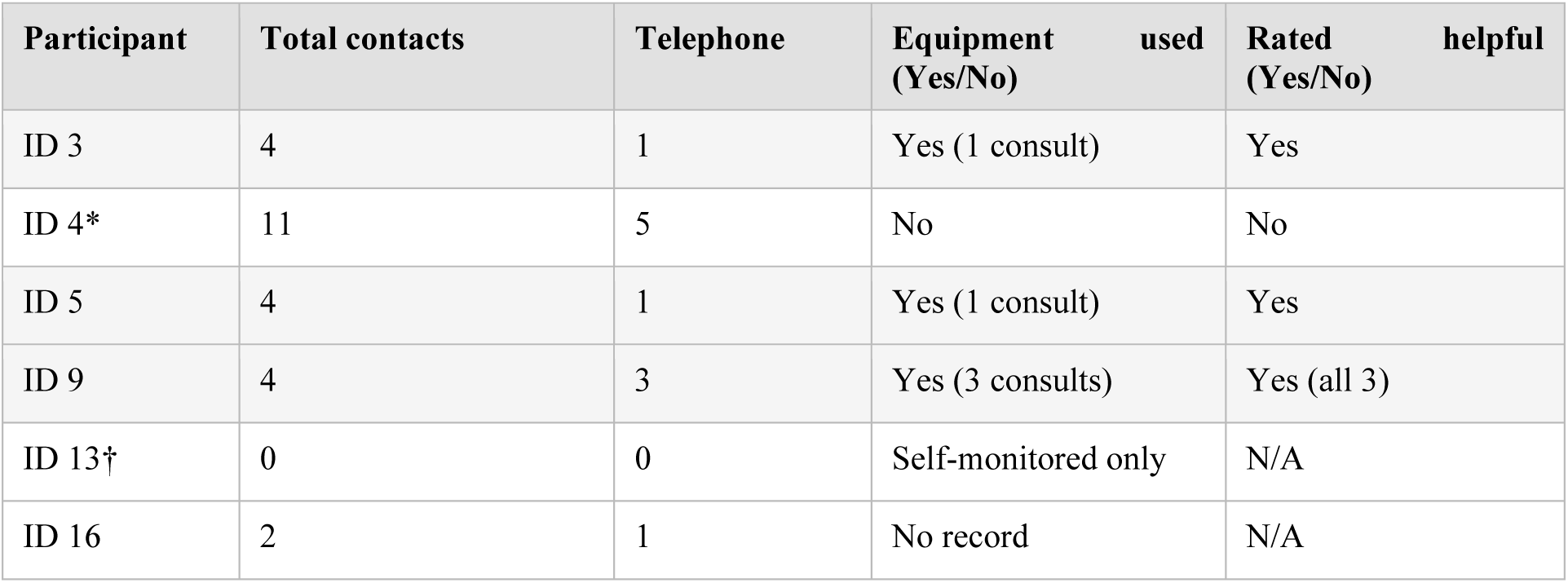
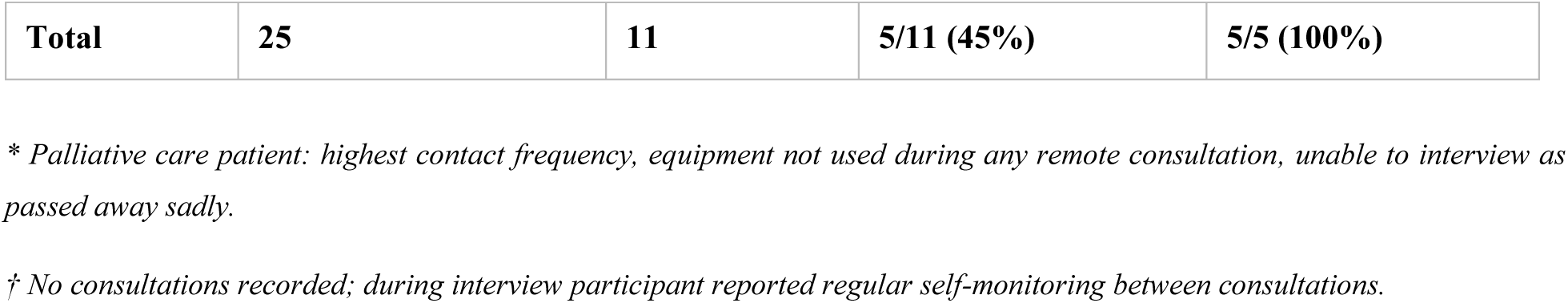
Equipment use per intervention group participant.

**Table 4.** Summary of Framework Analysis themes.

| Theme | Subthemes | Illustrative quote |
| --- | --- | --- |
| Facilitators of equipment use | Ease of use, improved access, self-monitoring, reassurance, portability | "Having the numbers when I phoned the surgery... they can't argue with the numbers" |
| Barriers to equipment use | Written instructions insufficient, medical terminology, GP non-engagement, labelling gaps | "The GP did not rely on my readings... I told them I had it but they didn't ask me to use it" |
| GP perspectives: in favour | Patient selection, useful clinical information, improved decision-making | GPs noted objective parameters supported remote triage and reassurance |
| GP perspectives: concerns | Increased workload, patient anxiety, lack of protocol, limited resources | "This will make patients more anxious... more appointments with borderline readings" |
| Suggestions for improvement | Video demonstrations, in-person training, terminology change ('wellness box') | "Consider calling it health equipment or well-being equipment... medical can be scary" |

One participant (ID 13) had zero primary care consultations during the six-month period but reported regular self-monitoring during their qualitative interview. The palliative care participant (ID 4), who had the highest number of contacts overall (n=11), did not use the equipment box during any of their five remote consultations. One participant (ID 16) had one remote consultation but no record of equipment use was found.

### Qualitative Findings

#### Patient Perspectives

Framework Analysis of semi-structured patient interviews identified five overarching themes. Facilitators of equipment use included ease of use (particularly for participants already familiar with one or more devices), a sense of improved access to GP services, reassurance gained from self-monitoring, portability of devices, and the ability to share objective readings during consultations. As one participant described, having numerical readings when phoning the surgery was a significant advantage: “They can’t argue with the numbers” providing a more objective basis for triage decisions than symptom descriptions alone.

Barriers identified included inadequate written instructions for some devices (particularly the blood pressure monitor), use of the word “medical” in the equipment name (perceived as anxiety-inducing), lack of engagement from GPs in requesting readings during consultations, and the absence of clear labelling on the clinical system to identify equipment holders. One participant noted that despite having taken readings before each telephone consultation, their GP did not ask for them, highlighting a systemic gap in protocol.

Participants made several suggestions for improvement, including the provision of demonstration videos alongside written instructions, the option for an in-person equipment demonstration at the surgery, and a terminology change from “Medical Self-Assessment Box” to “Wellbeing Box” or “Health Monitoring Equipment” to reduce the association with illness.

#### GP Perspectives

Framework Analysis of GP interviews identified three overarching themes. First, GPs perceived clear clinical value in patient-generated observations, particularly for respiratory and cardiovascular presentations, enabling safer remote triage and more confident decision-making. Objective parameters such as oxygen saturation, pulse, and blood pressure were reported to support reassurance, guide escalation thresholds, and occasionally prevent unnecessary face-to-face house visits.

Second, GPs highlighted the dual impact on patient experience: many patients gained reassurance and a sense of control from self-monitoring, yet a minority risked becoming anxious or over-monitoring, potentially increasing contacts with borderline readings.

Third, GPs emphasised important operational and implementation challenges, including variability in device accuracy, absence of clear coding or flagging for equipment holders in practice systems, and the need for defined monitoring schedules, patient guidance, and clinician protocols. Across interviews, GPs supported the intervention in principle but stressed that careful patient selection and clearer workflows would be essential for safe and sustainable implementation.

#### Integration of Findings

Integrating quantitative and qualitative findings, equipment non-use was explained by clinical context (palliative care, lack of remote consultations) rather than unwillingness. Qualitative data provided explanatory depth for the quantitative patterns observed, particularly the variability in equipment use across the intervention group. The consistent rating of the equipment as helpful when used, combined with patient-reported themes of improved access and reassurance, supports the feasibility of the intervention in appropriate patient groups.

#### Sample Size Calculation for Definitive Trial

Based on this feasibility study, an intervention uptake rate of approximately 50% was observed with excellent retention. For the definitive trial, assuming a primary outcome of the proportion of telemedicine consultations supported by use of the Medical Self-Assessment Box, and assuming a conservative usage rate of 25% in the control group and 50% in the intervention group, a total sample size of 130 participants (65 per group) would provide 80% power to detect this difference at a two-sided 5% significance level. Allowing for 10% attrition, the target sample size for a definitive trial would be 144 participants.

## 1. Discussion

### Main Findings

This mixed-methods feasibility study demonstrates that the ORCHARD intervention, providing medically vulnerable rural patients with a Medical Self-Assessment Box for use during telemedicine consultations, is feasible and acceptable when clinically appropriate. Recruitment was challenging, with only 15% of invited patients enrolling, reflecting the practical and regulatory difficulties of conducting feasibility trials in the post-pandemic primary care environment. However, retention was excellent at 100%, and all recruited participants accepted randomisation.

Among intervention participants, 50% used the equipment during at least one remote consultation, and equipment was used in 45% of telephone consultations where use was possible. Importantly, when used, the equipment was rated helpful in 100% of instances by patients. Non-use was largely explained by clinical context: the palliative care participant had the most consultations but appropriately did not use the equipment, and one participant self-monitored without having any recorded consultations. The intervention group had 31% fewer total primary care contacts than the control group, though the small sample size means this observation cannot be interpreted causally.

All remote consultations were conducted by telephone. The complete absence of video consultations during the study period is notable and may reflect both patient preference and the practice’s telemedicine infrastructure at the time of the study.

### Comparison with Existing Literature

These findings align with existing evidence on the value of home monitoring in supporting remote primary care decision-making. National policy initiatives in Scotland, including Connect Me, support home blood pressure and oxygen monitoring for specific conditions, and the ORCHARD findings extend this evidence to a broader medically vulnerable population in a rural context. The GP perspectives in this study echo findings from Greenhalgh and colleagues regarding the importance of the wider sociotechnical system in which remote care tools are embedded [13]: devices alone are insufficient without clear protocols, patient selection guidance, and clinician engagement.

The absence of video consultations contrasts with evidence that video consulting can reduce travel burden for rural patients [3,4] and suggests that implementation of video-based telemedicine in rural Scotland requires further attention. This may also reflect a need for direct research to inform rural and remote digital health policy.

### Strengths and Limitations

The key strength of this study is its mixed-methods design, which allowed quantitative feasibility data to be contextualised and explained by qualitative findings. The use of Framework Analysis, a structured and transparent method, enhances the credibility of the qualitative findings. The study also demonstrates that it is feasible to recruit medically vulnerable rural patients, randomise them, and collect meaningful data within a primary care setting.

Important limitations include the small sample size, which was substantially below the recruitment target of 40. A second invitation round may have improved recruitment. Participants were not clearly labelled in the clinical system as equipment holders, meaning GPs across in-hours and out-of-hours services were not reliably aware of which patients had the equipment. This is a significant implementation gap that must be addressed before a definitive trial. The palliative care participant could not be interviewed, limiting qualitative data from the highest-contact participant. The study was conducted at a single centre, limiting generalisability. SF-12 norm-based scoring was not possible without the licensed QualityMetric algorithm, limiting comparison with published norms.

### Implications for Policy and Practice

This study highlights the potential for self-monitoring medical equipment to support decision-making in remote primary care consultations, particularly for rural patients with complex health needs. However, technical literacy, clinical motivation, and clear implementation pathways must be addressed. Specifically, a definitive trial should incorporate: a clear clinical coding system to identify equipment holders; GP-facing protocols specifying when and how to request readings; a revised training package combining written instructions with demonstration videos; and consideration of terminology changes to reduce the anxiety associated with the word “medical.”

The complete absence of video consultations in this study underlines the need for direct acknowledgement of rural telemedicine challenges in national digital health policy across the four nations of the UK. The upcoming national digital plans represent an opportunity to ensure that the needs of rural and medically vulnerable populations are explicitly recognised.

## 5. Conclusion

The ORCHARD feasibility study demonstrates that providing medically vulnerable rural patients with a Medical Self-Assessment Box to support remote primary care consultations is feasible and acceptable when clinically appropriate. When patients self-monitor and report objective vital signs during teleconsultations, this information can meaningfully support clinical decision-making. A randomised controlled trial to evaluate the effectiveness of this intervention at scale is warranted. Future research should prioritise enhanced recruitment strategies, improved clinical system labelling of equipment holders, structured GP engagement protocols, and multimodal training materials. Telemedicine interventions tailored to rural populations offer significant promise for addressing persistent healthcare access inequities but must be rigorously tested through pragmatic, adequately powered trials.

## Author’s contribution

RA, PM made substantial contributions to the conception and design of the study, MN .SN contributed to data analysis and interpretation and was involved in manuscript drafting.All authors read and approved the final manuscript.

## Data Availability

All data produced in the present study are available upon reasonable request to the authors

## Acknowledgements

The authors gratefully acknowledge the time and commitment of those patient participants who gave their time to be interviewed in the study. We also acknowledge the contribution of, NHS Grampian, to the day to day running of the study.. This project was supported by a new investigator grant from

## REC reference

22/SW/0151 | RAS ID: 310499

## Trial registration

ClinicalTrials.gov Identifier: NCT05732922 Registered date: 16/02/2023

## Authors contributions

PM, RA led on the conception and design of the study. RA (Conceptualisation, funding acquisition, methodology, project administration, supervision and writing -review and editing).MN(participant recruitment, intervention application, data collection). SN(qualitative data collection , quantitative & qualitative data analysis, manuscript writing). All authors contributed to the interpretation of the findings (SN,RA,PM). SN drafted the first version of the manuscript with support from co-authors (PM,RA,MN) who provided a critical review. All authors (SN, RA,MN, PM) have approved the final version that has been submitted for publication.

## Acknowledgements

We are grateful to Co-I Finlayson for purposive sampling.Amanda Cardy, for help with organising payments to GP practice for GP interviews

## Target journal

Journal of Telemedicine and Telecare

## Appendices

### Appendix 1 ORCHARD Feasibility Study Flow Diagram

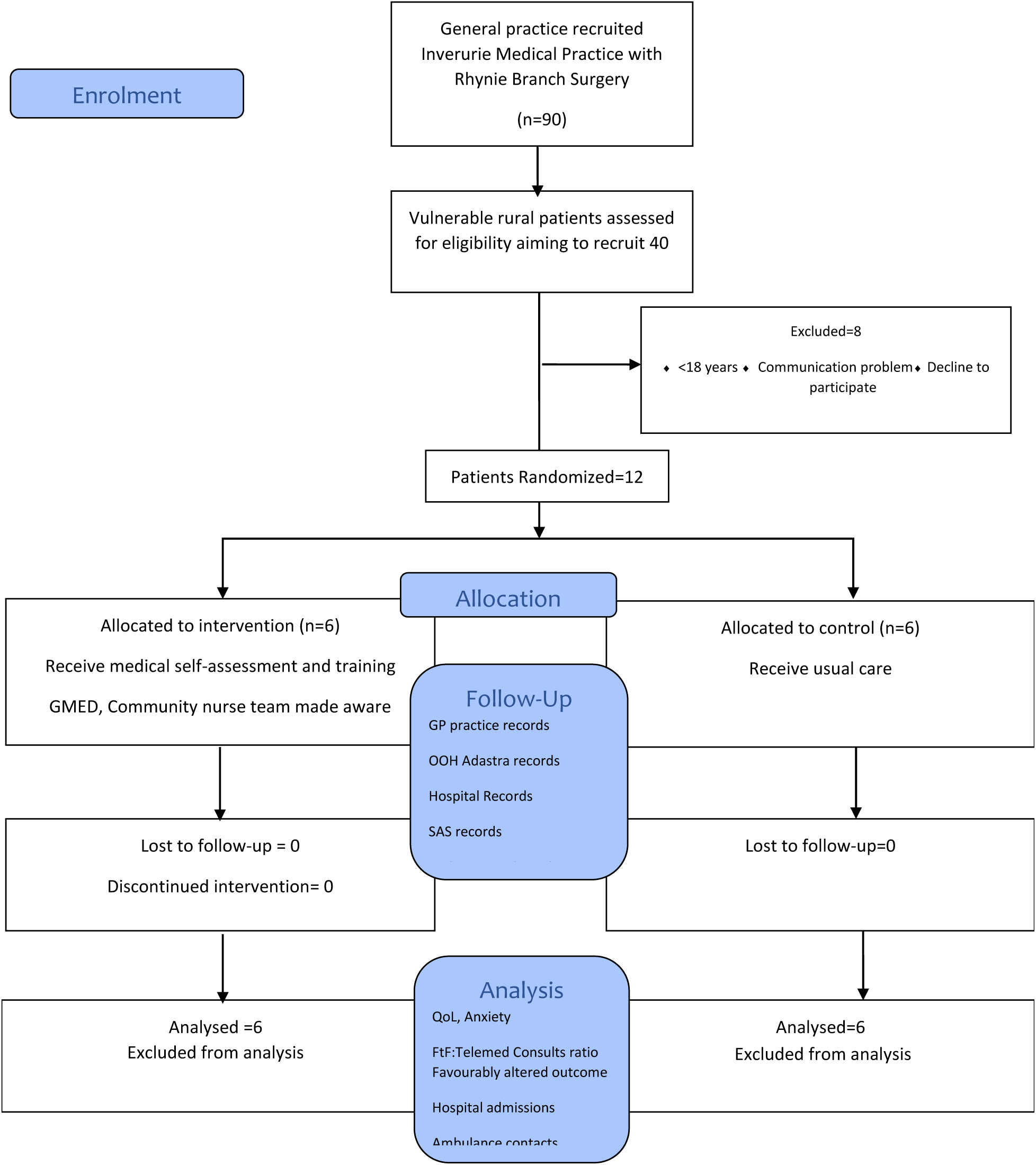

### APPENIDIX 2 – Criteria for shielded and at-risk patients

The Health Protection Scotland guidance and search criteria issued to Scottish Practice to identify shielded patients during the COVID-19 pandemic can be viewed at https://www.hps.scot.nhs.uk/web-resources-container/covid-19-search-criteria-for-highest-risk-patients-for-shielding/

People falling into the highest clinical risk group and eligible for shielding include:

1. Solid organ transplant recipients
2. People with specific cancers:

- people with cancer and are having chemotherapy
- people with lung cancer and are having radical radiotherapy
- people with cancers of the blood or bone marrow such as leukaemia, lymphoma or myeloma who are at any stage of treatment
- people having immunotherapy or other continuing antibody treatments for cancer
- people having other targeted cancer treatments which can affect the immune system, such as protein kinase inhibitors or PARP inhibitors
- people who have had bone marrow or stem cell transplants in the last 6 months, or who are still taking immunosuppression drugs
3. People with severe respiratory conditions including all cystic fibrosis, severe asthma and severe COPD
4. People with rare diseases and inborn errors of metabolism that significantly increase the risk of infections (such as SCID, homozygous sickle cell)
5. People on immunosuppression therapies sufficient to significantly increase risk of infection
6. Women who are pregnant with significant heart disease, congenital or acquired People falling into the next category of high or increased clinical risk include:

- are 70 or older
- are pregnant
- have a learning disability
- have a lung condition that’s not severe (such as asthma, COPD, emphysema or bronchitis)
- have heart disease (such as heart failure)
- have high blood pressure (hypertension)
- have diabetes
- have chronic kidney disease
- have liver disease (such as hepatitis)
- have a condition affecting your brain or nerves (such as Parkinson’s disease, motor neurone disease, multiple sclerosis, or cerebral palsy)
- have a problem with your spleen or have had your spleen removed
- have a condition that means you have a high risk of getting infections (such as HIV, lupus or scleroderma)
- are taking medicine that can affect your immune system (such as low doses of steroids)
- are very obese (a BMI of 40 or above)

See - https://www.nhs.uk/conditions/coronavirus-covid-19/people-at-higher-risk-from-coronavirus/whos-at-higher-risk-from-coronavirus/

### APPENDIX 3: Detailed Feasibility Questions

1. Are GPs, other healthcare professionals and practices willing to participate in the study?
2. Can data collected from practices and the GMED Out-Of-Hours Service be gathered to support the trial application and inform a detailed power calculation?
3. Can rural cancer patients be readily identified in searches of GP practice records?
4. Will rural cancer patients be willing to be recruited to a study of a Medical-Self-Assessment-Box?
5. Can the number and nature of daytime and out-of-hours consultations by recruited patients in the six-months preceding the study be readily determined from primary care-held medical records?
6. Are recruited rural cancer patients willing to be randomised to a trial of a Medical-Self-Assessment-Box?
7. Can a Medical-Self-Assessment-Box and instruction materials be distributed to recruited rural cancer patients?
8. Can rural cancer patients, randomized to receive a Medical-Self-Assessment-Box, use it effectively?
9. Which technical and practical problems arise when rural cancer patients are using a Medical-Self-Assessment-Box?
10. Can we flag participating patients effectively within their GP practice and to the GMED out-of-hours service via the Emergency Care Summary?
11. Will recruited patients complete a baseline and outcome questionnaire measuring their quality of life (SF-36) and psychological well-being PHQ4?
12. Will participating patients inform the trial team when they have used the Medical-Self-Assessment-Box?
13. Can we search GP Practice primary care-held and GMED Out-Of-Hours records twice weekly and identify encounters when the Medical-Self-Assessment-Box has been used?
14. Will GPs and Patients complete a brief electronic survey sent to them to gather data about consults when the Medical-Self-Assessment-Box was used?
15. Can we collect data on the number and nature of daytime primary care consultations by recruited patients?
16. Can we collect data on the number and nature of Out-Of-Hours by recruited patients?
17. Can we work out the ratio of Telemedicine:Face To Face consultations in primary care?
18. Can we collect appropriate data for a subsequent cost-effectiveness analysis?
19. Will patients and GPs agree to participate in qualitative interviews about their experiences?
20. Can we collect data on Scottish Ambulance Service Accident and Emergency attendances and hospital admissions for recruited patients?

### APPENDIX 4

#### Patient interview topic guides

Thinking about the last six months:

What are your thoughts about taking part in the study?

Prompt what was good about it?
Prompt- what was not so good?

Have you been regularly using the equipment when having a telemedicine consultation? Tell me how have you got on using the equipment?

Ease of use:
How easy did you find it to use?
Were you able to follow the instructions that accompanied it? Was it useful to give your GP some extra information?
Do you think the equipment is catering to all users?
Information:
Were the instructions for use accurate?
Were the instructions for use easy to understand? Were the instructions for use concisely written?
Was the language used on the instructions at an appropriate level (i.e. no jargon )? Were there any spelling or grammatical errors?
Are there sources for the information given (for example link to manufacturer’s website)?
Support:
Was there adequate access to support? Did you e-mail or call anyone?
Problems:
Did you have any problems/ technical difficulties with the equipment?
Did anything prevent you from using/ getting the most out of the equipment? Were there any aspects of the equipment design that were confusing?

Tell me about your use of medical equipment in general?

Do you use any medical equipment other than what we gave you? If So what do you use?
Tell me about how you monitored your blood pressure, blood gases, temperature, lung volume before we gave you this equipment?
Did you check yourself for any changes( regularly or just occasionally)?
Do you think that using equipment to report the accuracy of your condition to your GP would have been beneficial to you before the study? (have you changed your mind now?)

Were there any occasions when you were concerned about your health?

Tell us about what happened?

Thinking about your experience of using this equipment?

What were the best aspects of using the equipment?
What were the worst aspects of using the equipment?
Do you have any suggestions for how we could make our study better?
Does the use of equipment fulfil its purpose?

Does using like equipment give you more Peace of Mind about giving a more accurate picture of your health condition to your GP?

Would you recommend the use of this equipment?

Were there any unintended consequences from the use of the equipment?

If yes what were they?

### Appnedix 5

#### GP interview topic guides

Thinking about the last six months

What are your thoughts about this study?

Prompt what was good about it?
Prompt what was not so good?

Have you been regularly asking for readings when having a telemedicine consultation? How did your patients get on using the equipment?

Ease of use:
How easy did they find it to use?
Were they able to follow the instructions that accompanied it? Was it useful to give you some extra information?
Do you think the equipment is catering to all users? Next line Information:
Were the instructions for use accurate?
Were the instructions for use easy to understand? Were the instructions for use concisely written?
Was the language used on the instructions at an appropriate level (i.e.no jargon? Were there any spelling or grammatical errors?
Are there sources for information given (e.g. link to manufacturers website?) Support:
Was there adequate access to support? Did you e-mail or call anyone?
Problems:
Did your patients have any problems/ technical difficulties with the equipment? Did anything prevent you from using/ getting the most out of the equipment?
Were there any aspects of the equipment design that your patients found confusing?

Tell me about how your patients monitored the blood pressure, blood gases, temperature, lung volume before we gave them this equipment?

Do you think that your patients using equipment to report the accuracy of their condition during a telemedicine consultation would have been beneficial to you before the study(Have you changed your mind now?)

Were there any occasions when you would concerned about the patients health?

Tell us about what happened?

Thinking about your experience of using this equipment?

What were the best aspects of your patients’ use of equipment?
What were the worst aspects of your patients’ use of equipment?
Do you have any suggestions on how we could make our study make our study better?
Does the use of equipment fulfil its purpose?

Does your patients’ use of the equipment give you more Peace of Mind about giving more accurate advice to them?

Would you recommend the use of this equipment?

Were there any unintended consequences from the use of the equipment?

If yes,what were they?

- Do you recommend the use of this equipment to your patients? If yes what do you say?
- What do you think about the length and time that is involved in the use of equipment?
- Do you think it is feasible for your patients to self monitor?
- Do you think patients will find it easy or difficult to perform these measurements? Why?
- Have your patiennts ever suggested to you that they find self monitoring worriyng or reassuring? Which method, home or clinic , do you think evaluates the measured parameters more reliably?
- What do you feel are the main issues to consider when thinking about self monitoring at home?

## References

1. Scottish Government. COVID-19 Search criteria for highest risk patients for shielding. https://www.hps.scot.nhs.uk (Accessed 13 May 2020)

2. Turner M, Fielding S, Ong Y, et al. A cancer geography paradox? Poorer cancer outcomes with longer travelling times to healthcare facilities despite prompter diagnosis and treatment. Brit J Cancer 2017;117:439–449.

3. Ronis SD, McConnochie KM, Wang H, Wood NE. Urban telemedicine enables equity in access to acute illness care. Telemedicine and e-Health 2017;23(2):105–12.

4. Khairat S, Haithcoat T, Liu S, et al. Advancing health equity and access using telemedicine: a geospatial assessment. J Am Med Inform Assoc 2019;26(8-9):796–805.

5. Greenhalgh T, Koh GCH, Car J. Covid-19: a remote assessment in primary care. BMJ 2020;368:m1182.

6. Kaplan A, Hauptman R. The virtual COPD visit in the COVID-19 pandemic. Canadian Family Physician 2020.

7. Bugge C, et al. A process for Decision-making after Pilot and feasibility Trials (ADePT). Trials 2013;14:353.

8. Maxwell M, et al. The impact of rurality on patient experience and diagnostic pathway intervals in Scotland’s cancer patients. National Cancer Diagnosis Audit, 2023.

9. Murchie P, Adam R, Khor WL, et al. Impact of geography on Scottish cancer diagnoses in primary care. Cancer Epidemiol 2020;66:101720.

10. Levin KA. Urban-rural differences in self-reported limiting long-term illness in Scotland. J Public Health Med 2003;25(4):295–302.

11. Brundisini F, Giacomini M, DeJean D, et al. Chronic disease patients’ experiences with accessing health care in rural and remote areas: a systematic review. Ont Health Technol Assess Ser 2013;13(15):1–33.

12. Bhatia S, Landier W, Paskett ED, et al. Rural-Urban Disparities in Cancer Outcomes. J Natl Cancer Inst 2022;114(7):940–952.

13. Greenhalgh T, Wherton J, Papoutsi C, et al. Beyond Adoption: A New Framework for Theorizing and Evaluating Nonadoption, Abandonment, and Challenges to the Scale-Up, Spread, and Sustainability of Health and Care Technologies. J Med Internet Res 2017;19(11):e367.

14. Greenhalgh T, Koh GCH, Car J. Covid-19: a remote assessment in primary care. BMJ 2020;368:m1182.

15. National Institute for Health and Care Excellence (NICE). Covid-19 rapid guideline: managing suspected or confirmed pneumonia in adults in the community. NICE guideline NG165. 2020.

16. Scottish Government. Care in the Digital Age: Delivery Plan 2025–2026. Edinburgh: Scottish Government; 2024.

